# Quantifying age-associated differences in the durability of Shingrix-induced immunity using mathematical modelling

**DOI:** 10.64898/2026.11.10.26367539

**Authors:** Terry Easlick, Guy De La Rosa, Soren Gantt, Haijing Qin, Lisa R. Shelton, David A. G. Skibinski, Morgan Craig

## Abstract

Understanding the durability of vaccine-induced immunity is critical for evaluating long-term protection and vaccine scheduling, particularly in aging populations where immunosenescence may alter immune response dynamics. In this study, we used a novel mathematical modelling pipeline to quantify age-related differences in the contraction and persistence phases of gE-specific CD4+ T cell and anti-gE antibody responses following Shingrix vaccination. Specifically, we developed a semi-mechanistic model to expand virtual populations that captured inter-individual variability in peak concentrations and waning dynamics. To compare immune responses across age groups, we introduced a time varying, age-related immune advantage metric that measures the probability of a randomly selected younger individual exhibiting a stronger immune response than one from an older group. By evaluating this metric across multiple simulated virtual populations, our results demonstrate clear and sustained age-related differences in both gE-specific CD4+ T cells and anti-gE antibody responses, with younger age groups consistently exhibiting higher responses. Given these age-related differences, we used our model to explore alternative vaccine schedules in older age groups. These exploratory simulations suggest that booster doses administered between 6 and 12 months following the second dose may mitigate age-related differences in antibody dynamics. This work highlights the use of mechanistic mathematical modelling to disentangle age-associated differences in vaccine outcomes and lead to designing of plausible improvement in age-specific vaccine schedules.

## INTRODUCTION

Herpes zoster (HZ), commonly known as shingles, is an infectious disease caused by the reactivation of latent varicella-zoster virus (VZV). Primary exposure to VZV results in chickenpox, typically during childhood, after which the virus establishes lifelong latency.^1^ Reactivation events may subsequently occur years or decades later. Vaccination is the primary strategy for preventing HZ and its complications.

Shingrix is a recombinant glycoprotein E (gE) AS01B adjuvanted HZ vaccine (HZ/su) that is administered as a two-dose intramuscular series in adults aged 50 years and older to prevent HZ. Its efficacy and safety was demonstrated in the phase III ZOE-50/ZOSTER-006 randomized, placebo-controlled multicentre clinical trial.^2^ Broadly speaking, old age is associated with reduced immune responses to many vaccines and infections.^3–8^ Age-associated changes in vaccine-induced immunity are particularly important to consider since these differences may not only be present in the magnitude of the initial response but also the subsequent durability. In particular, Shingrix immunogenicity, vaccine efficacy and duration of protection have been found to be higher among younger adults than older adults.^9^ Long-term protection against HZ is particularly important, as age-related declines in immune control increase susceptibility to VZV reactivation, elevating the risk of HZ in older adults.^10^ Given the target population of Shingrix is adults 50 and older, it serves as a valuable case study in understanding the impacts of ageing on durability of immunity.

Understanding how cellular and humoral responses persist in ageing populations is crucial for the evaluation of vaccine strategies. Protection against HZ is likely primarily mediated by T cells.^11^ However, the kinetics of both cellular and humoral immune responses to gE induced by Shingrix may be important since both could be serve as measures of long-term vaccine-induced protection.^9^ Understanding the contraction and long-term persistence of antigen-specific CD4+ T cell and antibody responses is therefore valuable for evaluating vaccine durability and informing immunization strategies in these ageing populations. However, these processes are biologically complex and only partially observable experimentally. This complexity therefore motivates the use of mechanistic mathematical models to quantitatively characterize immune kinetics and infer underlying biological mechanisms. Mathematical modelling provides a useful framework for characterizing immune-response kinetics, including waning, persistence, and long-term maintenance dynamics that are difficult to measure directly.^12^ More broadly, model-informed approaches are increasingly used in drug and vaccine development to integrate diverse sources of evidence, support quantitative decision-making, and optimize development strategies throughout the development cycle.^13–16^ Previous studies have applied mechanistic ordinary differential equation (ODE) models to characterize vaccine-induced immune responses following HZ vaccination.^17,18^ In parallel, virtual population (VPOP) approaches have increasingly been used to represent inter-individual heterogeneity in mechanistic models.^19–21^

In this study, we developed a semi-mechanistic mathematical model to describe the long-term contraction and persistence dynamics following Shingrix vaccination. Using data from the ZOSTER-006 clinical trial^2^, we generated virtual patient populations for both gE-specific CD4+ T cell and anti-gE antibody responses to characterize age-dependent immune persistence. To compare immune persistence across age cohorts, we introduced a time-varying probabilistic metric of superiority defined as the probability that a randomly selected younger virtual individual exhibits a greater immune response than a randomly selected older virtual individual at a given timepoint. Using this metric, we showed how age-related differences in immune persistence can be quantified over time and used to evaluate the booster responses required to overcome these disadvantages. Thus, our framework provides a quantitative approach for assessing and optimizing age-specific vaccination strategies.

## MATERIALS and METHODS

### Data

To calibrate our model and expand virtual populations (VPOPs), we used data of both cellular and humoral responses from the GSK Clinical Study Report from the ZOSTER-006 trial^2^ in which participants were stratified into three age groups (50-59, 60-69, and 70+ years) and received vaccine doses at months 0 and 2. Measurements were collected prior to vaccination (baseline, month 0) and at multiple post-vaccination timepoints at months 3, 14, 26, and 38 (Figure 1). The gE-specific CD4+ T cell frequency (per 10^6^ cells) was quantified using intracellular cytokine staining with flow cytometry analysis, while anti-gE antibody concentrations were measured using enzyme-linked immunosorbent assay (ELISA).

**Figure 1.**
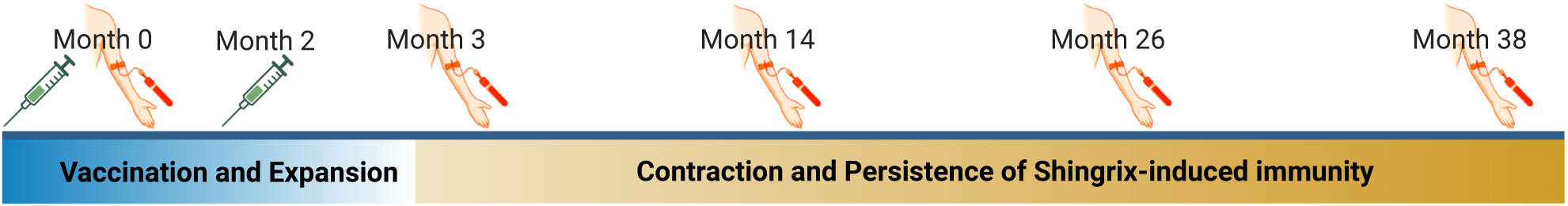
Dosing and sampling timeline. Participants received doses at month 0 and month 2. Peripheral blood mononuclear cell samples for gE-specific CD4+ and serum samples for anti-gE antibody concentrations were collected on months 0, 2, 3, 14, 26, and 38. The study’s entire timeline lasted slightly more than three years.

The available data consisted of aggregated summary statistics for gE-specific CD4+ T cell frequencies expressing at least two activation markers among IL-2, IFN-γ, TNF-α, and CD40L and anti-gE antibody ELISA concentrations (mIU/mL) rather than individual-level longitudinal measurements. Specifically, the available summaries included the number of participants (including missing participants), mean, standard deviation, minimum, median, quartiles, and maximum at each timepoint within each age cohort. Depending on the age-cohort and time-point, the number of participants with available gE-specific CD4+ data ranged from 45 to 63, while the number with available anti-gE antibody data ranged from 297 to 356. Hereafter, we used the conversion of 1 month = 30 days for modelling. Since the data provide measurements at the time of first dose (day 0) and approximately 30 days following the second dose (day 90), we assumed the day 90 observation corresponds to the first post-peak measurement. This is consistent with previous studies reporting that gE-specific CD4+ T cell frequencies peak approximately 7-14 days following vaccination.^22^ Additionally, prior studies indicate that antibody concentrations peak approximately one month following vaccination and as early as two weeks post-vaccination in elderly adults.^23,24^

It is important to note the gE-specific CD4+ and anti-gE antibody data were cohort-level rather than at the individual-level; thus, we considered the data for each separately. Additionally, the data for each age-cohort was handled independently of the other two age-cohorts.

### Mathematical model of gE-specific CD4+ T cell and anti-gE antibody responses following Shingrix vaccination

Following vaccination, cellular and humoral immune responses typically display three distinct phases: 1) initial expansion, 2) contraction, and 3) long-term persistence. During the expansion phase, antigen-specific CD4+ T cells undergo rapid proliferation before reaching their peak and subsequently progressively declining cell number (contraction). These phases ultimately give rise to a smaller, long-lived memory CD4+ T cell population that contributes to sustained immune protection. In parallel, antigen-specific B cells differentiate into antibody-secreting plasma cells, producing a peak in circulating antibody concentrations followed by progressive waning as short-lived plasma cells decay. Long-term antibody persistence is maintained through long-lived plasma cells and antigen-specific memory B-cells.

To describe the contraction and persistence of both gE-specific CD4+ T cell and anti-gE antibody responses following Shingrix vaccination, we developed a semi-mechanistic mathematical model. Let *x*(*t*) be the total abundance of either the gE-specific CD4+ T cells, *T*, or anti-gE antibody concentrations, *Ab*, and *t*_1_ denote the post-peak observation time. As our primary objective was to predict long-term immune persistence, we did not explicitly model proliferation dynamics. Then, for both CD4+ T cells and antibody concentrations, the contraction and persistence phase dynamics are described by:

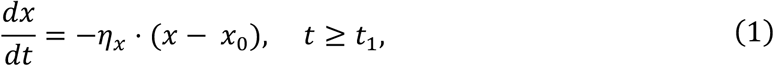

where *η_X_* is the decay rate (1/day) of *x*, and *x*_0_ denotes the baseline abundance, subject to the initial condition

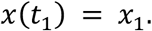

A schematic description of the model is provided in Figure 2.

**Figure 2.**
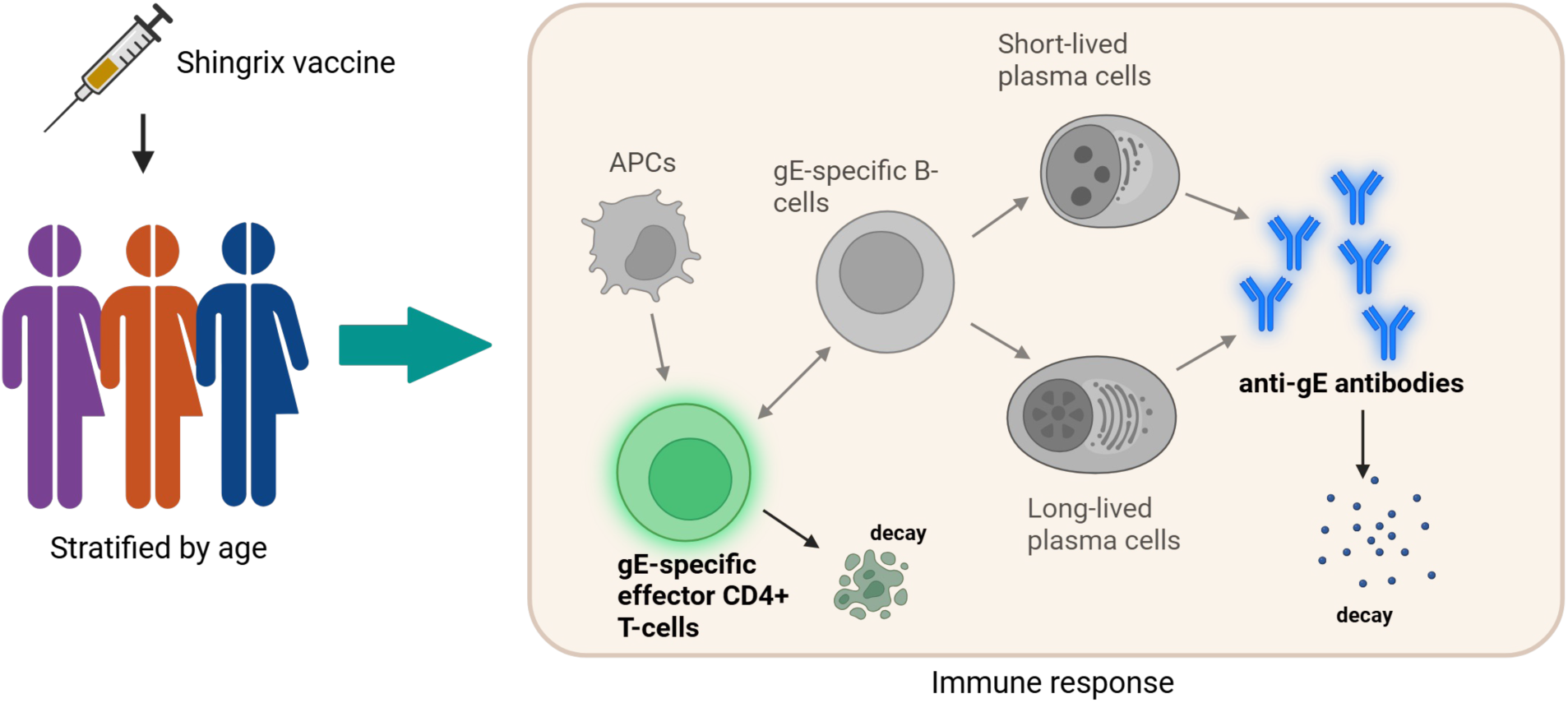
Model schematic for immune response post Shingrix vaccination. Participants were partitioned into three age cohorts consisting of 50-5S, C0-CS, and 70+ years of age. They were administered two doses of Shingrix C0 days apart. To describe their immune dynamics following vaccination, we used the model described in the beige shaded box. Upon vaccination, APCs stimulate CD4+ T cells to become antigen (gE) specific. These gE-specific CD4+ cells aid in the creation of gE-specific B cells, ultimately leading to the creation of plasma cells and finally anti-gE antibodies. Coloured compartments: modelled immune components. Greyed compartments: known immune components not described in our model.

### Virtual Population Generation and Parameter Estimation

To account for inter-individual variability and for each age-cohort, the model parameters *X*_0_, *X*_1_ and *η_x_* were assumed to follow lognormal distributions

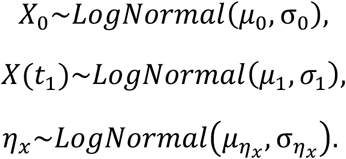

This assumption is common practice for strictly positive and right-skewed biological response data ^22,23^.

For each age-cohort, cohort specific distributions for *x*_0_ and *x*_1_ were reconstructed directly from the reported median and interquartile range using known distributional relationships under the lognormal assumption. Note that as the data non-zero baselines for both gE-specific CD4+ T cells and anti-gE antibody concentrations, we used the baseline value as a lower bound for the long-term immunity (hence, the inclusion of *x*_0_ in Eq. (1)).

An initial estimate for the decay parameter *η_x_* for each age-cohort was obtained by fitting Eq. (1) to the median reported data value. Unlike the distributions for *x*_0_ and *x*_1_, the distribution parameters *μ_ηx_* and σ*_ηx_* were obtained using an approximate version of trajectory-matching random-walk feasibility sampling (TM-RWFS) framework.^18^ Briefly, virtual populations were generated by sampling baseline and peak immune responses from the empirical distributions and decay rates from candidate log-normal distributions. Simulated longitudinal trajectories were compared with the observed cohort using an ABC discrepancy metric, and the decay-rate distribution was iteratively refined using a Metropolis–Hastings acceptance criterion. To preserve the observed cohort structure, we generated virtual populations with virtual cohort sizes approximately matching the corresponding study populations. Depending on the age cohort and observation time, the number of participants with available gE-specific CD4+ measurements ranged from 45–63; whereas the number with available anti-gE antibody measurements ranged from 297–356.

For proposed values of *μ_ηx_* and σ*_ηx_*, a virtual patient *i* in a given age-cohort was generated by sampling:

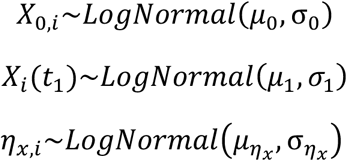

where *i* = 1, … , *N* with *N* chosen to approximately match the observed cohort size for the corresponding age-cohort and biomarker.

The mechanistic model given in Eq. (1) was then simulated for all virtual patients, and cohort-level summary statistics were calculated at each observation time. Simulated summary statistics were compared against the reported summary statistics, and candidate parameter distributions were accepted within the TM-RWFS framework when sufficient agreement was achieved. Repeated application of this procedure yielded an ensemble of accepted VPOPs.

### Statistical metric to quantify age-related differences

As the central question of our study was age-related immunological differences, we defined a novel statistical metric to predict immune persistence across age cohorts. This measure quantifies the probability that a randomly selected younger virtual individual exhibits a greater immune response than a randomly selected older virtual individual at a given timepoint, which we interpret as the immune *advantage* of one individual over another. Let *A* denote a virtual cohort that is younger relative to the virtual cohort *B*. The advantage probability between these two virtual populations at time *t* was defined as

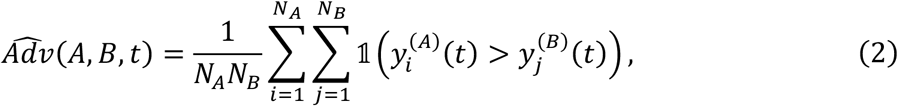

where *N_A_* and *N_B_* denote the numbers of virtual individuals in populations *A* and *B*, respectively, 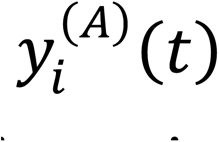 and 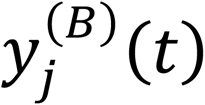 denote the respective corresponding simulated immune-response trajectories at time *t*, and 1 is an indicator function whose value is equal to 1 when the sampled immune response from population *A* exceeds that from population *B* and 0 otherwise. In the absence of a measurable advantage, 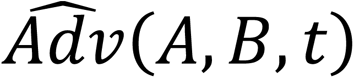 is expected to remain near the null threshold of 0.5. Deviations above this threshold indicate an immune response advantage for population *A*, whereas values below 0.5 indicate a relative disadvantage.

### Assessing robustness

To assess the robustness of age-group comparisons, the ABC-MCMC calibration procedure was repeated 100 times, generating 100 independent VPOPs. The immune advantage metric was evaluated separately within each VPOP.

## RESULTS

### Age-stratified virtual population reveals that gE-specific CD4+ T cell waning increases with age

To quantify age-related differences in the persistence of Shingrix-induced immunity, we created an age-stratified virtual population. For each age cohort within the VPOP, we generated the same number of virtual individuals as the number of available ZOSTER-006 trial observations available at the day 90. Using Eq. 1, we simulated the longitudinal dynamics of gE-specific CD4+ T cells following Shingrix vaccination. The model predicted trajectories show that all cohorts (i.e., 50-59, 60-69, and 70+) exhibited progressive contraction of vaccine-induced CD4+ T cell responses over time (Figure 3A). Although younger virtual individuals generally maintained higher long-term CD4+ T cell frequencies than older individuals, these differences were not readily apparent through visual inspection of the trajectories alone. This agrees with the ZOSTER-006 trial data.

**Figure 3.**
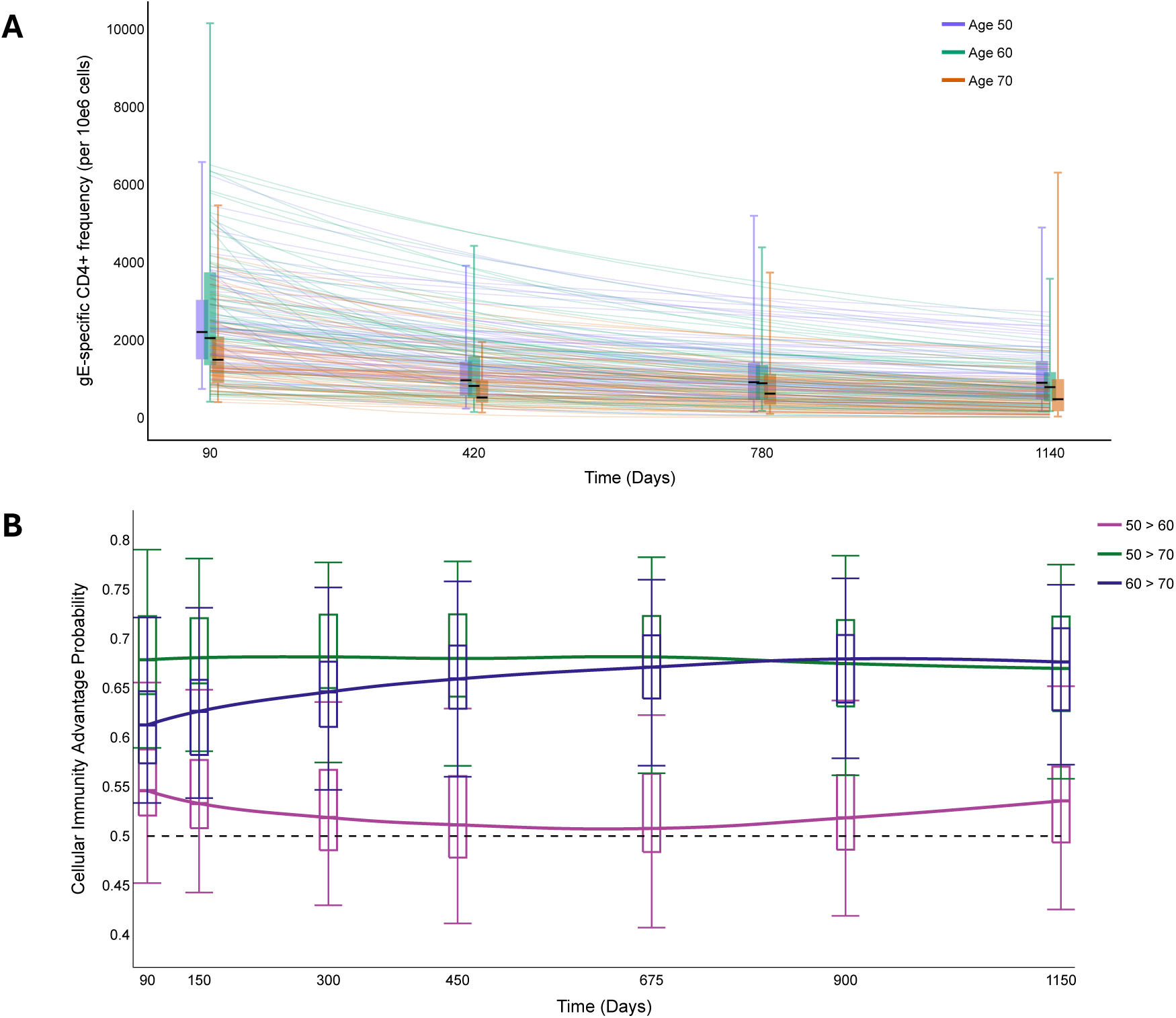
Virtual population analysis of gE-specific CD4+ T cell persistence dynamics across age cohorts following Shingrix vaccination. A) Simulated longitudinal persistence trajectories for virtual subjects in the 50-59, 60-69, and 70+ age cohorts. Vertical lines: ranges of reported data. Boxplots: medians and interquartile ranges in previously reported^2^ cohort summary statistics. Whiskers: min and max. B) Time-varying advantage probabilities comparing age-stratified virtual cohorts from 100 independently generated VPOPs. Values above 0.5 indicate a greater likelihood that the younger cohort exhibits a higher immune response than the older cohort at a given time point. Boxplots: medians and interquartile ranges. Whiskers: 95% prediction intervals.

To test for age-related differences in CD4+ count trajectories more directly, we evaluated the time-varying advantage probability (Figure 3B) between our age-stratified virtual cohorts. As explained in the methods, this metric, 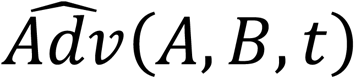, quantifies the immune advantage (here, the number of gE-specific CD4+ T cells) of one randomly selected virtual individual in a younger cohort (i.e., *A*) over a randomly selected virtual individual in an older cohort (i.e., *B*). Our results showed a persistent age-associated advantage in long-term gE-specific CD4+ T cell responses for younger cohorts, with the strongest effect found between the 50-59 versus 70+ cohorts and the smallest between the 50-59 versus 60-69 virtual cohorts where 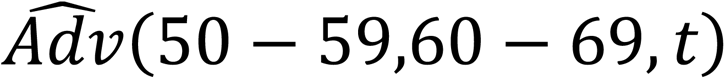 remained close to the null 0.5 threshold throughout the study period. Notably, the advantage probability for the 50-59 versus 70+ comparison remained relatively stable over time, whereas the advantage displayed by the 60-69 cohort relative to the 70+ cohort increased progressively throughout the persistence phase.

### Younger virtual individuals display an anti-gE antibody concentration advantage over older virtual individuals

The same analyses of the anti-gE antibody concentrations revealed similar patterns of age-related differences in immune persistence. As for the gE-specific CD4+ T cell frequencies, simulations of our semi-mechanistic model showed good agreement with the ZOSTER-006 trial data across the 50-59, 60-69, and 70+ virtual cohorts (Figure 4A), all of which exhibited substantial inter-individual heterogeneity. This resulted in considerable overlap among simulated trajectories throughout the observation period. Although visual differences between age groups were modest, younger age cohorts were found to generally maintain stronger long-term antibody responses than the oldest cohort.

**Figure 4.**
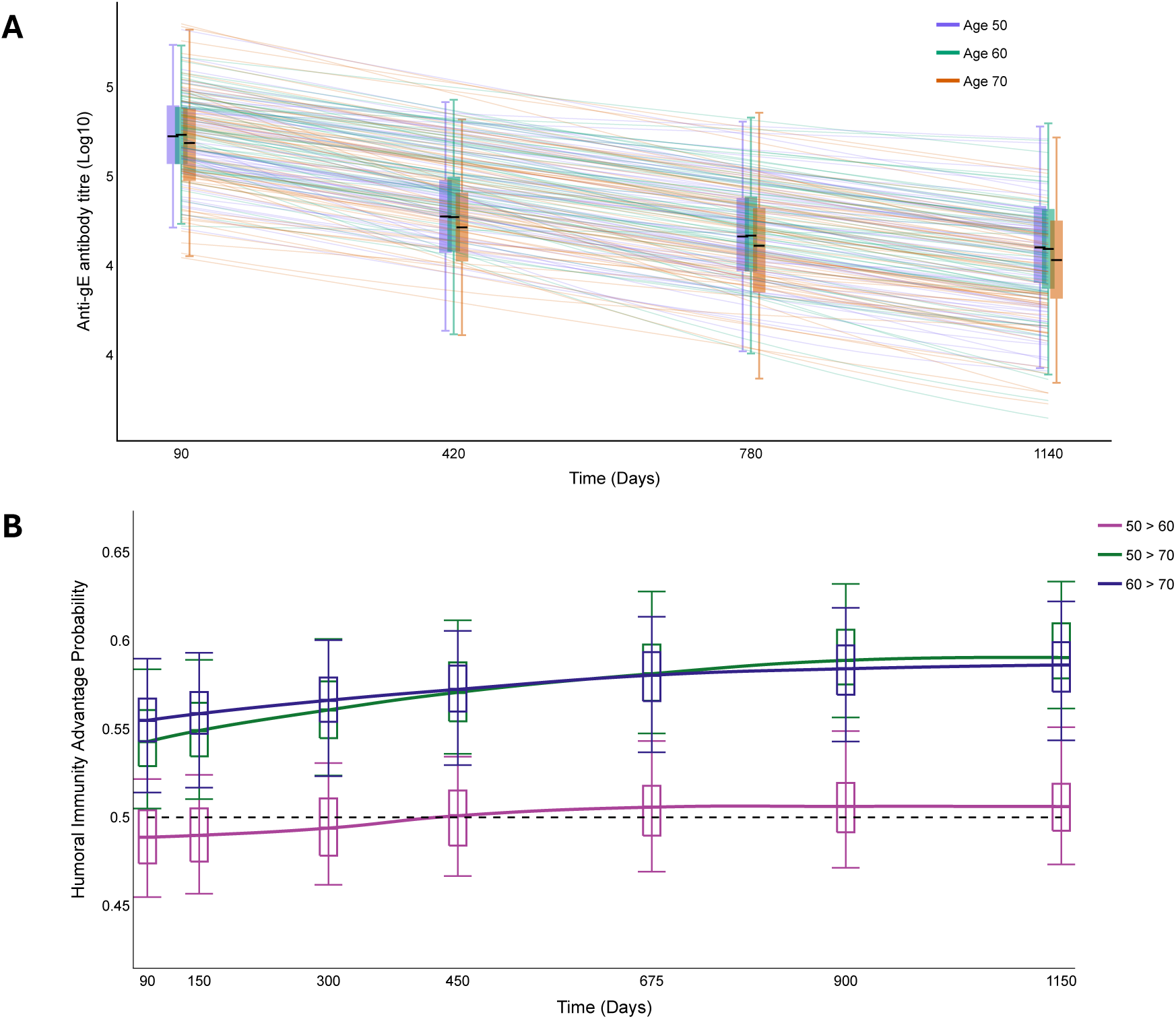
Virtual population analysis of anti-gE antibody concentration persistence dynamics across age cohorts following Shingrix vaccination. A) Simulated longitudinal persistence trajectories for virtual subjects in the 50-59, 60-69, and 70+ age cohorts in Log10 scale. Vertical lines: ranges of reported data. Boxplots: medians and interquartile ranges in previously reported cohort summary statistics.^2^ Whiskers: min and max. B) Time-varying advantage probabilities comparing age-stratified virtual cohorts from 100 independently generated VPOPs. Values above 0.5 indicate a greater likelihood that the younger cohort exhibits a higher immune response than the older cohort at a given time point. Boxplots: medians and interquartile ranges. Whiskers: 95% prediction intervals.

Thus, to check for age-related differences, we evaluated the advantage probability between age-stratified virtual populations (Figure 4B). These analyses also detected an advantage for younger cohorts (i.e., 50-59 versus 70+, and 60-69 versus 70+), both of which remained consistently above the null threshold of 0.5 across all observed follow-up times. Moreover, this advantage increased progressively over time, suggesting stronger long-term persistence in the younger age groups relative to the 70+ cohort. In contrast, the 50-59 versus 60-69 comparison remained close to the null threshold throughout the study period, indicating relatively minor differences between these two younger age groups. Together, these results may suggest that differences in anti-gE antibody concentrations in older individuals may be driven by differences in gE-specific CD4+ T cell waning detected using our model, or an unmodelled biological mechanism.^25^

### Age-associated differences in immune durability show signs of reduction with an additional booster for 70+ virtual cohort

Having identified persistent age-related disadvantages in the 70+ age cohort, we next explored whether these differences could be mitigated through the administration of an additional booster dose. Hence, using the VPOPs and parameters obtained as described above, we simulated an additional booster dose as an instantaneous increase in the immune response at a specified booster time, *t*_boost_. Let *α_x_* (unitless) represent the immunogenicity strength of the additional booster. We modelled the effects of a third dose by modulating the immunogenicity strength variable *α_x_* relative to the observed immunity level at day 90 (time *t*_1_):

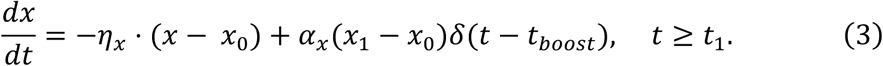

Third doses were simulated to occur either 180 or 365 days after the first Shingrix dose. For each virtual subject in the 70+ cohort, simulated gE-specific CD4+ T cell frequency and anti-gE antibody concentrations were evaluated at days 1180 and 1365, respectively, corresponding to 1000 days after administration of the third dose. As no third dose was simulated for the younger cohorts (50–59 and 60–69 years), the same biomarkers were evaluated at day 1060, corresponding to 1000 days after completion of the primary two-dose series. This ensured that comparisons were made at the same interval (1000 days) following the most recent vaccine dose. The advantage probability was then calculated by comparing the younger virtual cohorts with the boosted 70+ virtual cohort. Finally, we determined the minimum third-dose immunogenicity scaling factor required such that the median advantage probability crossed 0.5 or the scaling factor reached 1.0, whichever occurred first.

Our model predicted that the age-related advantages in both gE-specific CD4+ T cell frequencies and anti-gE antibody concentrations could be reduced or eliminated through an additional booster in the oldest, 70+ virtual cohort. This is indicated by the median advantage probability trending towards the 0.5 value as the second booster strength was increased. Our simulations further show the importance of planning the proper booster sizes relative to its timing (Figure 5). For example, a booster administered at day 180 required strength response less than that of one administered on day 365. However, in both scenarios the required strength of the booster approaches 100% relative to the cellular immunity at day 90 (time *t*_1_) to only begin cancelling out the age advantage of some younger virtual subjects.

**Figure 5.**
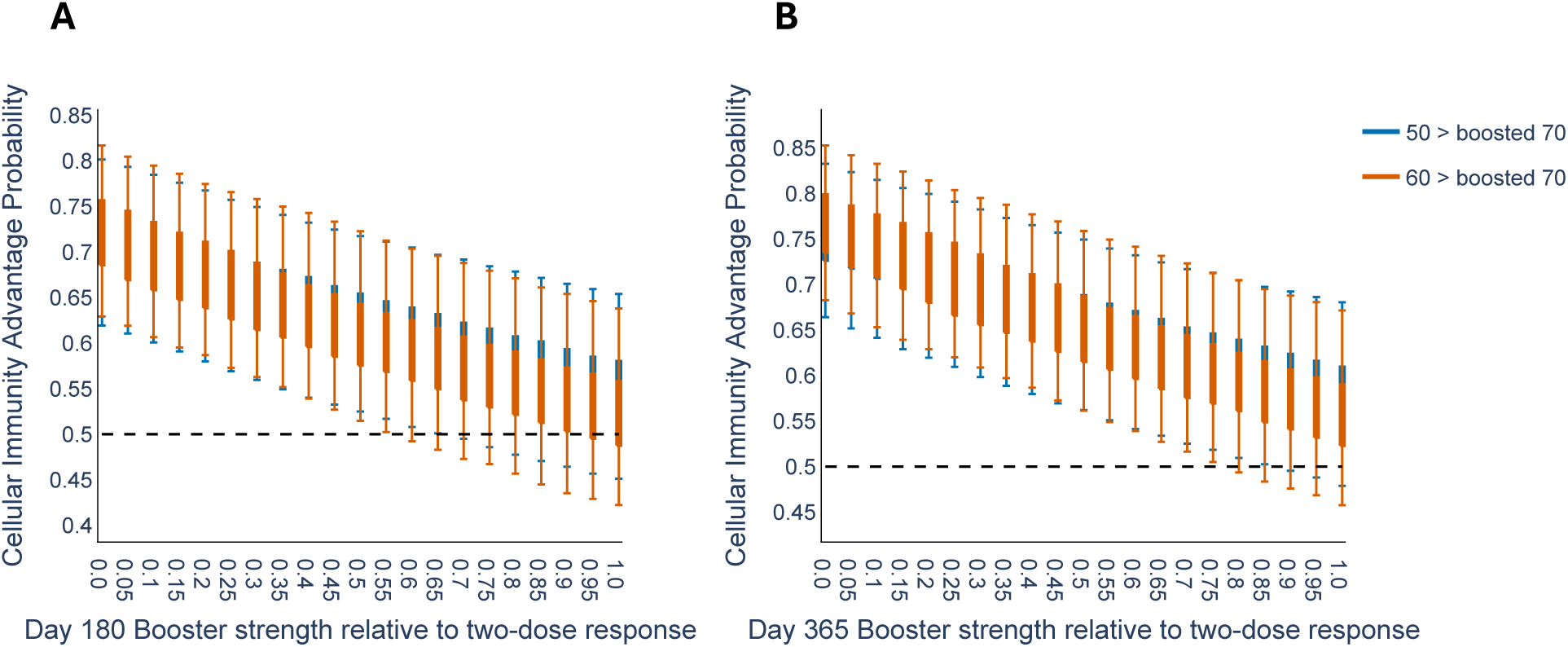
Impact of hypothetical booster-dose strength on age-related gE-specific CD4+ T cell advantage probabilities. Distributions in gE-specific CD4+ T cell advantage probabilities from 100 independently generated VPOPs between younger (i.e., 50-59 and 60-69) virtual cohorts and oldest (i.e., 70+) virtual cohort after the administration of a second booster given A) 180 days or B) 365 days after the first Shingrix dose. The earlier an additional booster was given, the larger its dose immunogenicity strength was required to be to offset the age-related advantage in gE-specific CD4+ T cell frequencies. A) The first quartile of the advantage probability crosses the null threshold of 0.5 only when the booster response is approximately equivalent to the immunity level at day 90. B) A booster dose at one year indicates even less capability to overcome cellular immunity advantage held by the younger age-cohorts. A-B) Boxplots: medians and interquartile ranges. Whiskers: 95% prediction intervals.

In contrast, weaker booster responses were better able to offset the age-related disadvantage in anti-gE antibody persistence. For the day 180 booster, the median advantage probability crossed the null threshold when the booster response was approximately 50-55% of the day 90 immune response. To accomplish the same for the day 365 booster it would need to achieve approximately 75-80% of the day 90 humoral immunity (Figure 6B). The higher booster threshold required for gE-specific CD4+ T cell responses indicates that the age-related disadvantage, as quantified by the advantage metric, is more difficult to overcome for cellular immunity than for humoral immunity, supporting the idea that CD4+ T cells may drive age-related disparities in immune responses.

**Figure 6.**
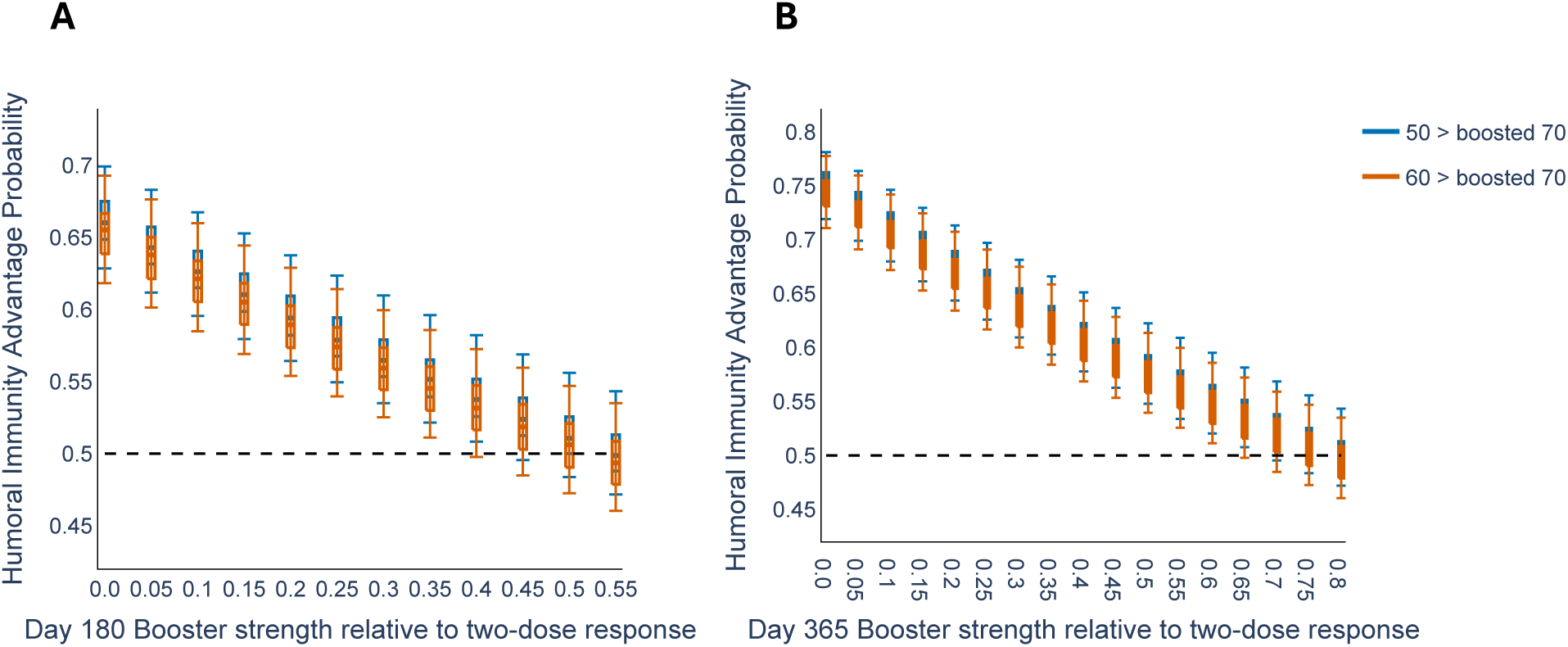
Advantage probability for anti-gE antibody after 70+ age-cohort booster. Distributions in anti-gE antibody concentrations advantage probabilities from 100 independently generated VPOPs between younger (i.e., 50-59 and 60-69) virtual cohorts and oldest (i.e., 70+) virtual cohort after the administration of a second booster given A) 180 days or B) 365 days after the first Shingrix dose. A) Booster dose administered at 180 days post-first dose required a booster response approaching 55% of the original day 90 response level to nullify the median advantage probability. B) A booster administered 365 days after the first Shingrix dose required a response approximately 80% of the original day 90 response to nullify the median advantage probability. A-B) Boxplots: medians and interquartile ranges. Whiskers: 95% prediction intervals.

## DISCUSSION

Our results identify persistent age-associated differences in both cellular and humoral immune response following Shingrix vaccination. Despite inter-individual heterogeneity in the age groups, the individuals in the 70+ virtual cohort consistently exhibited lower long-term gE-specific CD4+ T cell and anti-gE antibody responses than younger virtual cohorts. Importantly, these age-associated difference were not identical between the cellular and humoral immunity.

In this study, we developed a virtual population modelling framework to investigate age-related differences in the long-term persistence of Shingrix-induced immunity. Though we found significant overlap in simulated longitudinal trajectories for both gE-specific CD4+ T cells and anti-gE antibodies due to a high degree of inter-individual heterogeneity across age-stratified virtual cohorts, our novel advantage-probability metric detected systematic differences between age groups. Indeed, across both cellular and humoral responses, the estimated advantage probabilities remained consistently above the null threshold of 0.5 for all comparisons involving the 70+ virtual cohort, suggesting measurable waning of immune strength with older age. This highlights one of the primary strengths of the proposed approach: rather than relying solely on comparisons of average trajectories or visually distinguishable cohort separation, the immune advantage metric quantifies the chance that one age cohort exhibits a stronger persistent immune response than another cohort.

As we detected important differences in the strength of the immune response in virtual individuals over 70 years-of-age, we hypothesized that it would be possible to “rescue” the immune response of older individuals to recover the response exhibited by those between 50 and 69 years-old. To test this, we studied how strong a second booster dose administered 180 and 365 days after the first dose would need to be to nullify the advantage displayed by 50-to-69-year-olds when measured 1000 days after the most recent Shingrix administration. We found that a second booster dose administered to adults aged 70 and older six months or one year after the first can result in greater persistence of immunity as that observed in younger adults. Furthermore, our model predicted that a second booster administered at 180 days after the first Shingrix dose would need to generate a weaker response relative to the original two-dose series than a booster administered at one year to nullify the median advantage probability. With respect to gE-specific CD4+ T cell frequencies, the booster dose strength would need to be quite strong, approaching 100% of the immune response present at day 90, in particular, which we assumed was already post-peak response. However, when we consider anti-gE antibody concentration response, the day 180 dose was predicted to need around 50-55% strength as compared to the day 90 values, indicating that the age-related decline in antibody response can perhaps be overcome. The day 365 booster dose strength to overcome anti-gE antibody concentration advantages in younger virtual individuals required approximately 75-80% of the day 90 values. Overall, the strength required to begin reducing humoral immunity advantages were smaller than that for the cellular immunity advantages. Moreover, our results indicate it is more difficult to mitigate age-related advantages in cellular immunity than for humoral immunity. Whether this pattern can be demonstrated empirically in the clinic and whether it generalizes to other vaccines warrants further investigation. We note that because no clinical third-dose data were available, these analyses should be interpreted as exploratory threshold analyses rather than calibrated predictions of booster immunogenicity.

Our study is limited by the available data, which consisted primarily of baseline measurements together with longitudinal observations following administration of the second vaccine dose. Thus, the expansion phase of the immune response was not observable with sufficient temporal resolution to support detailed quantitative modelling. Although the qualitative behaviour of the expansion phase, including rapid immune activation and progression toward peak response, is biologically well understood, sparse sampling during this interval limits the ability to reliably estimate parameters governing expansion kinetics. Age-related differences in early immune activation and peak-response timing are likely to exist; however, these effects could not be robustly quantified within the present framework. Further, there was a notable difference in sample sizes available for the two immune-response measurements: whereas anti-gE antibody analyses were based on substantially larger cohort sizes of approximately 300–350 participants per age-cohort, the gE-specific CD4+ T cell analyses were only approximately 45–63 participants per age-cohort. Consequently, we would expect the virtual populations to provide more stable estimates of population-level variability and persistence anti-gE antibody dynamics. Nevertheless, the CD4+ T cell cohorts remained of moderate size and were sufficient to identify consistent age-related trends across independently generated virtual populations. Therefore, while the smaller CD4+ T cell sample sizes may contribute to greater uncertainty in the simulated trajectories and advantage probabilities, they are unlikely to qualitatively alter the primary conclusions regarding age-associated differences in immune persistence. Although the aggregated data did not allow for the estimation of correlations between model parameters, the independent sampling approach provided a practical means of capturing inter-individual heterogeneity while reproducing the observed cohort-level immune response dynamics. Lastly, biologically, both gE-specific CD4+ T cell and anti-gE antibody persistence arise from interactions of short-lived and long-lived immune populations, including effector and memory CD4+ T cells, plasmablasts, long-lived plasma cells, and memory B cells. Though we initially explored more complex model formulations intended to separately represent these components, the available data was insufficiently granular to reliably identify multiple decay timescales. Thus, we intentionally simplified the model used here by using a single decay process rather than explicitly modelling multiple immune subpopulations to describe the contraction and persistence dynamics of both immune responses to ensure parameter identifiability given the available data resolution. Future work could extend the present framework, particularly if more densely sampled longitudinal immunogenicity data during the expansion phase immediately following vaccination were available. This would permit explicit modelling of activation and proliferation dynamics together with more detailed characterization of age-related differences in early immune kinetics. Additional mechanistic complexity could also be incorporated through multi-compartment models describing short-lived and long-lived immune populations separately. Furthermore, integration of individual-level longitudinal measurements would allow more rigorous estimation of inter-individual variability and potentially improve the realism of the generated virtual populations.

Despite these limitations, this study nonetheless demonstrates that mechanistic VPOP approaches provide a useful framework for understanding and quantifying long-term vaccine-induced immune persistence in ageing heterogeneous populations. Moreover, demonstrating the usefulness in deriving novel biological insight that can be used to guide pre- and post-clinical vaccine development. Indeed, even under limited observational data, the combination of mechanistic modelling, virtual population generation, and probabilistic comparison metrics enables quantitative characterization of age-associated immune durability that may be difficult to assess directly from empirical cohort summaries alone.

Our results strongly underline the need to understand how ageing impact cellular and humoral immune response differently. Further, our study highlights the importance of considering differing dosing schedules for different age-groups and suggests that older adults may benefit from an additional booster dose to help alleviate possible impacts of immunosenescence on their long-term immunity. These results are particularly important against the backdrop of aging populations, a decade past the approval of Shingrix. Ultimately, this work motivates the continued study of age-related immune effects through collaborative efforts integrating predictive mathematical modelling. In our work, Shingrix served as a case study; however, our results make clear the need for further investigation into the age-associated vaccine-induced immune durability across other vaccines used in ageing populations: Is the age-associated loss of immune durability observed here also present in other vaccinations? Are the age-associated differences in cellular and humoral immunity similarly reflected in other vaccine outcomes?

## DECLARATIONS

### Ethics approval and consent to participate

Not applicable. These results used publicly available data from a previously conducted clinical trial.

### Consent for publication

Not applicable. No individual data was utilized.

### Availability of data and materials

The data analyzed in this study are available from the GSK clinical study register at https://www.gsk-studyregister.com/trials/110390. Formatted data and code are included on the study’s Github https://github.com/Craig-Lab/QuantAgeAssocVaccineDurability.

### Authors’ contributions

TE, SG, HQ, LRS, DAGS, and MC conceived and designed the research. TE, LRS, DAG, and MC performed the research. TE analyzed the results. TE, GDLR, SG, HQ, LRS, DAGS, and MC wrote and edited the manuscript.

## Data Availability

All data used in this study is available at the GSK clinical trials study registry.

https://www.gsk-studyregister.com/

## Acknowledgements

Not applicable.

## References

1 Cohen, J. I. Herpes Zoster. New England Journal of Medicine 369, 255–263 (2013). doi:10.1056/NEJMcp1302674

2. Study to evaluate efficacy, safety and immunogenicity of GSK Biologicals’ Herpes Zoster (HZ) vaccine GSK1437173A in adults aged 50 years and older, <https://www.gsk-studyregister.com/trials/110390> (2014).

3 Lal, H. et al. Efficacy of an adjuvanted herpes zoster subunit vaccine in older adults. N Engl J Med 372, 2087–2096 (2015). 10.1056/NEJMoa1501184

4 Cunningham, A. L. et al. Immune Responses to a Recombinant Glycoprotein E Herpes Zoster Vaccine in Adults Aged 50 Years or Older. The Journal of Infectious Diseases 217, 1750–1760 (2018). 10.1093/infdis/jiy095

5 Chen, J., Deng, J. C. & Goldstein, D. R. How aging impacts vaccine efficacy: known molecular and cellular mechanisms and future directions. Trends in Molecular Medicine 28, 1100–1111 (2022). 10.1016/j.molmed.2022.09.008

6 Wagner, A. et al. Age-related differences in humoral and cellular immune responses after primary immunisation: indications for stratified vaccination schedules. Sci Rep 8, 9825 (2018). 10.1038/s41598-018-28111-8

7 Doherty, T. M., Weinberger, B., Didierlaurent, A. & Lambert, P.-H. Age-related changes in the immune system and challenges for the development of age-specific vaccines. Annals of Medicine 57 (2025). 10.1080/07853890.2025.2477300

8 Lord, J. M. The effect of aging of the immune system on vaccination responses. Human Vaccines & Immunotherapeutics 9, 1364–1367 (2014). 10.4161/hv.24696

9 Strezova, A. et al. Final analysis of the ZOE-LTFU trial to 11 years post-vaccination: efficacy of the adjuvanted recombinant zoster vaccine against herpes zoster and related complications. eClinicalMedicine 83 (2025). 10.1016/j.eclinm.2025.103241

10 Chiavarini, M. et al. Vaccination Against Herpes Zoster in Adults: Current Strategies in European Union Countries. Vaccines 13 (2025). 10.3390/vaccines13101073

11 Weinberg, A. & Levin, M. J. in *Varicella-zoster Virus Current Topics in Microbiology and Immunology* Ch. Chapter 31, 341–357 (2010).

12 Garcia-Fogeda, I., Besbassi, H., Larivière, Y., Ogunjimi, B., Abrams, S. & Hens, N. Within-host modeling to measure dynamics of antibody responses after natural infection or vaccination: A systematic review. Vaccine 41, 3701–3709 (2023). 10.1016/j.vaccine.2023.04.030

13 Bai, J. P. F. et al. The case for model-informed vaccine development. Vaccine 92 (2026). 10.1016/j.vaccine.2026.128780

14 Desikan, R., Germani, M., van der Graaf, P. H. & Magee, M. A Quantitative Clinical Pharmacology-Based Framework For Model-Informed Vaccine Development. Journal of Pharmaceutical Sciences 113, 22–32 (2024). 10.1016/j.xphs.2023.10.043

15 Zhou, J. et al. Model-Informed Drug Development Applications and Opportunities in mRNA-LNP Therapeutics. Clinical Pharmacology & Therapeutics 118, 1329–1339 (2025). 10.1002/cpt.3641

16 Sahasrabudhe, V., Nicholas, T., Nucci, G., Musante, C. J. & Corrigan, B. Impact of Model-Informed Drug Development on Drug Development Cycle Times and Clinical Trial Cost. Clinical Pharmacology & Therapeutics 118, 378–385 (2025). 10.1002/cpt.3636

17 Keersmaekers, N., Ogunjimi, B., Van Damme, P., Beutels, P. & Hens, N. An ODE-based mixed modelling approach for B- and T-cell dynamics induced by Varicella-Zoster Virus vaccines in adults shows higher T-cell proliferation with Shingrix than with Varilrix. Vaccine 37, 2537–2553 (2019). 10.1016/j.vaccine.2019.03.075

18 Besbassi, H. et al. Modeling antibody dynamics following herpes zoster indicates that higher varicella-zoster virus viremia generates more VZV-specific antibodies. Front Immunol 14, 1104605 (2023). 10.3389/fimmu.2023.1104605

19 Craig, M., Gevertz, J. L., Kareva, I. & Wilkie, K. P. A practical guide for the generation of model-based virtual clinical trials. Front Syst Biol 3, 1174647 (2023). 10.3389/fsysb.2023.1174647

20 Beigmohammadi, F., Weaver, J. J. A., Hegarty-Cremer, S., Jeynes-Smith, C., Smith, A. M. & Craig, M. Trajectory matching-random walk feasibility sampling for simulating heterogeneous dynamics in mechanistic models. bioRxiv (2025). 10.1101/2025.09.26.678800

21 Allen, R. J., Rieger, T. R. & Musante, C. J. Efficient Generation and Selection of Virtual Populations in Quantitative Systems Pharmacology Models. CPT: Pharmacometrics & Systems Pharmacology 5, 140–146 (2016). 10.1002/psp4.12063

22 Kim, C., Fang, F., Weyand, C. M. & Goronzy, J. J. The life cycle of a T cell after vaccination - where does immune ageing strike? Clin Exp Immunol 187, 71–81 (2017). 10.1111/cei.12829

23 Lane, A., Quach, H. Q., Ovsyannikova, I. G., Kennedy, R. B., Ross, T. M. & Einav, T. High-resolution antibody dynamics following influenza vaccination reveal predominantly weak responses as well as infrequent but durable immunity across the 2014-2022 seasons. Vaccine 64, 127677 (2025). 10.1016/j.vaccine.2025.127677

24 Gross, P. A., Russo, C., Teplitzky, M., Dran, S., Cataruozolo, P. & Munk, G. Time to peak serum antibody response to influenza vaccine in the elderly. Clinical Diagnostic Laboratory Immunology 3, 361–362 (1996). 10.1128/cdli.3.3.361-362.1996

25 Deng, X., Farhang-Sardroodi, S. & Craig, M. Predicting age-related determinants of heterogeneous outcomes to COVID-19 mRNA vaccines through mathematical modelling. *preprint* (2025). 10.1101/2025.02.14.25322308

